# Epidemiological characteristics of deaths from COVID-19: an analysis at almost three months of the first confirmed case in Peru

**DOI:** 10.1101/2020.11.05.20226639

**Authors:** Willy Ramos, Juan Arrasco, Jhony A. De La Cruz-Vargas, Luis Ordóñez, María Vargas, Yovana Seclén, Miguel Luna, Nadia Guerrero, José Medina, Isabel Sandoval, Edith Solís, Manuel Loayza

## Abstract

**OBJECTIVE:** To determine the epidemiological characteristics of deaths from COVID-19 in Peru from March 28 to May 21, 2020, 85 days after the report of the first confirmed case.

**MATERIAL AND METHODS:** Case series type study. Deaths from various sources were investigated, including the COVID-19 Epidemiological Surveillance and the National System of Deaths. Deaths that met the definition of a confirmed case and had a positive (reactive) result of RT-PCR or rapid test were considered for the analysis. From these sources, epidemiological variables were obtained and a time analysis was performed, defining as the pre-hospital time the time from the onset of symptoms to the date of hospitalization and hospital time from the date of hospitalization to the date of death.

**RESULTS:** 3,244 confirmed deaths were included in the study. Deaths were more frequent in males (71.8%), elders (68.3%), residents of the department of Lima (41.8%), and others from the coast (37.7%). In 81.3% of cases, the death occurred in a public hospital, 16.0% died at home, shelter, penitentiary institution, public highway, or in transit to a hospital, and 31.1% had some comorbidity.

Statistical difference was observed in pre-hospital time according to age group (p <0.001) and gender (p = 0.037), being significantly higher in adults, elders, and women. There was a statistically significant difference in hospital time according to geographic area, showing a significantly shorter time in the departments of the coast (p <0.001) and Andean region (p = 0.014) compared to Lima. The cases that were seen in private clinics (p = 0.001) survived longer than those seen in public hospitals.

**CONCLUSION:** Deaths from COVID-19 occur mainly in male, elders, on the coast, with considerable deaths at home, in shelters, penitentiaries, public roads, or in transit to a hospital. Pre-hospital time is affected by age group and gender; while, hospital time is also influenced by the region of origin and the health care provider.

## INTRODUCTION

The current COVID-19 pandemic reported since December 2019 constitutes the biggest public health problem in decades. To this scenario are added the diversity of economic, social, and demographic aspects, and the different responses of health systems to identify and provide medical care to affected people.^1-3^

The evolution of the pandemic, in terms of incidence, mortality, and speed of expansion, is heterogeneous, with differences between countries and even between regions of the same country. Some studies report differences in the clinical presentation and severity of the disease; Likewise, a part of those infected will require health care, including hospitalization, therefore, the structure and capacity of health systems are important for the impact of the pandemic.^1,4,5^

Since the introduction of the pandemic in Peru, epidemiological surveillance was implemented including the notification of deaths, which is a useful tool to measure the impact of COVID-19 and mortality is the most important final indicator in pandemic circumstances. The number of deaths from COVID-19 can be influenced by the diagnostic testing strategy, health system responsiveness and the social behavior of the population, the dynamic situation between countries, among other factors.^3,6,7^

In these circumstances, the analysis of the epidemiological characteristics of deaths from COVID-19 including the pre-hospital and hospitalization time intervals (elapsed days) could provide valuable information related to the barriers of accessibility, capacity, health system responsiveness, course and severity of the disease. These allow to understand the behavior, evolution, and trends of the health emergency in Peru and other Latin American countries.^3.8^

The Republic of Peru has a population of 32 495 500 inhabitants distributed in 24 departments and a constitutional province. It is grouped into three natural regions: the coast between the coast, the Andean region, located on the mountain range of the Andes, and the jungle that encompasses the Peruvian Amazon^9^. Lima is the capital of Peru and it is located on the coast with a population that exceeds 10 million inhabitants.^10^

According to the aging index, Peru is ranked number 48 out of 96 positions in the Global Aging Index and has a life expectancy of 76.5 years. The Encuestas Demográficas y de Salud Familiar (ENDES) as well as the health analyses show that the Peruvian population has a health profile dominated by noncommunicable diseases such as high blood pressure, cardiovascular diseases, cancer, and diabetes mellitus, comorbidities that constitute factors risk for COVID-19. People aged 80 years or older have the highest comorbidity rates (67.6%).^11-14^

In May 20, 2020, the Centro Nacional de Epidemiología, Prevención y Control de Enfermedades del Ministerio de Salud del Perú had reported 108,769 confirmed cases of COVID-19 and 3,148 deaths with lethality of 2.89%. At that time, there was a high demand for people who required health services compared to insufficient supply, which caused the health establishments to be overwhelmed by a large number of cases while the Ministry of Health tried to organize the response with the available resources.^15^

By the end of August 2020, Peru became the country with the highest mortality rate from COVID-19 in the world, reaching 89.43 deaths per one hundred thousand inhabitants, so the analysis of the characteristics of the deaths is of national and international interest^16^. The objective of this research was to determine the epidemiological characteristics of deaths from COVID-19 in Peru from March 28 to May 21, 2020, 85 days after the report of the first confirmed case.

## METHODOLOGY

Report of case series type. The database of the epidemiological surveillance of the coronavirus disease was revised, including the deaths investigated by the personnel of the Epidemiology Offices of the notifying health facilities at the national level. The quality control and monitoring of the notification were carried out by Epidemiology personnel at the national level, sector workers assigned to each region, and Regional Health Directorates.

Confirmed deaths from COVID-19 were considered for analysis; it included only those with a positive (reactive) RT-PCR result, rapid tests, or both that were notified between March 28 and May 21, 2020. Deaths not attributable to COVID-19 were excluded from the study; that is, deaths caused by entities or processes in which there was no evidence that COVID-19 infection changed the course of the disease. The sources of information on deaths from COVID-19 were as follows.

− Sistema Nacional de Defunciones en Línea (SINADEF)
− Database of RT-PCR test results (Netlab) and/or rapid tests (SISCOVID).
− Clinical histories of hospitalized cases.
− If complete information on the death was not available from these sources, telephone communication was made with the epidemiologists of the regions, who in some cases performed a verbal autopsy.

From these sources, epidemiological variables such as age, gender, department of residence (probable place of infection), date of onset of symptoms, date of hospitalization, date of death, place where the death occurred, health provider institution, and diagnosis of comorbidities. In the case of comorbidities, diabetes mellitus, arterial hypertension, cardiovascular diseases, cancer, chronic kidney disease, obesity, chronic pulmonary and bronchial diseases were considered.

A time analysis was performed, the pre-hospital time was defined as the time from the onset of symptoms to the date of hospitalization, and time in hospital as the time from the date of hospitalization to the date of death. Finally, it was analyzed whether the pre-hospital and hospital times showed differences according to the epidemiological characteristics evaluated. The analysis was guided under the assumption that a longer pre-hospital time indicates a delay in reaching a health facility and that a longer hospital time is an indicator of a higher survival of the cases.

The statistical analysis was carried out with the statistical program SPSS version 25 for Windows. Descriptive statistics were performed based on obtaining frequencies, percentages, measures of central tendency, and dispersion. Because the variables pre-hospital time and hospital time did not present a normal distribution (Kolmogorov-Smirnov test) and the variances were heterogeneous (Levene’s test), the bivariate analysis was performed with non-parametric tests such as the Kruskal-Wallis test and the Mann-Whitney U test. In the case of the Kruskal-Wallis Test, a posteriori analysis was performed with the Dunn-Bonferroni Test. The calculations were performed with a confidence level of 95%.

## RESULTS

During the study period, 3,263 deaths confirmed by COVID-19 were registered, of which 11 were excluded because the death occurred due to incidental causes (acute myocardial infarction, ruptured cerebral aneurysm, diabetic ketoacidosis, tuberculous meningoencephalitis, sepsis of dermal focus and perforated appendicitis) and 8 due to incomplete data, 3244 deaths being available for analysis. Of the 3,244 deaths, 95.7% corresponded to pneumonia; while the remaining 4.3% corresponded to home deaths of cases with very fast-evolving respiratory symptoms not explained by another disease.

### EPIDEMIOLOGICAL CHARACTERISTICS

As of May 22, 2020, the number of deaths confirmed by COVID-19 in Peru showed an upward trend; however, there was a drop in the number of deaths in the last week of the study due to the duration of the investigate or delay of confirmatory results of RT-PCR tests, which were later regularized (Graph 1).

**FIGURE 1:**
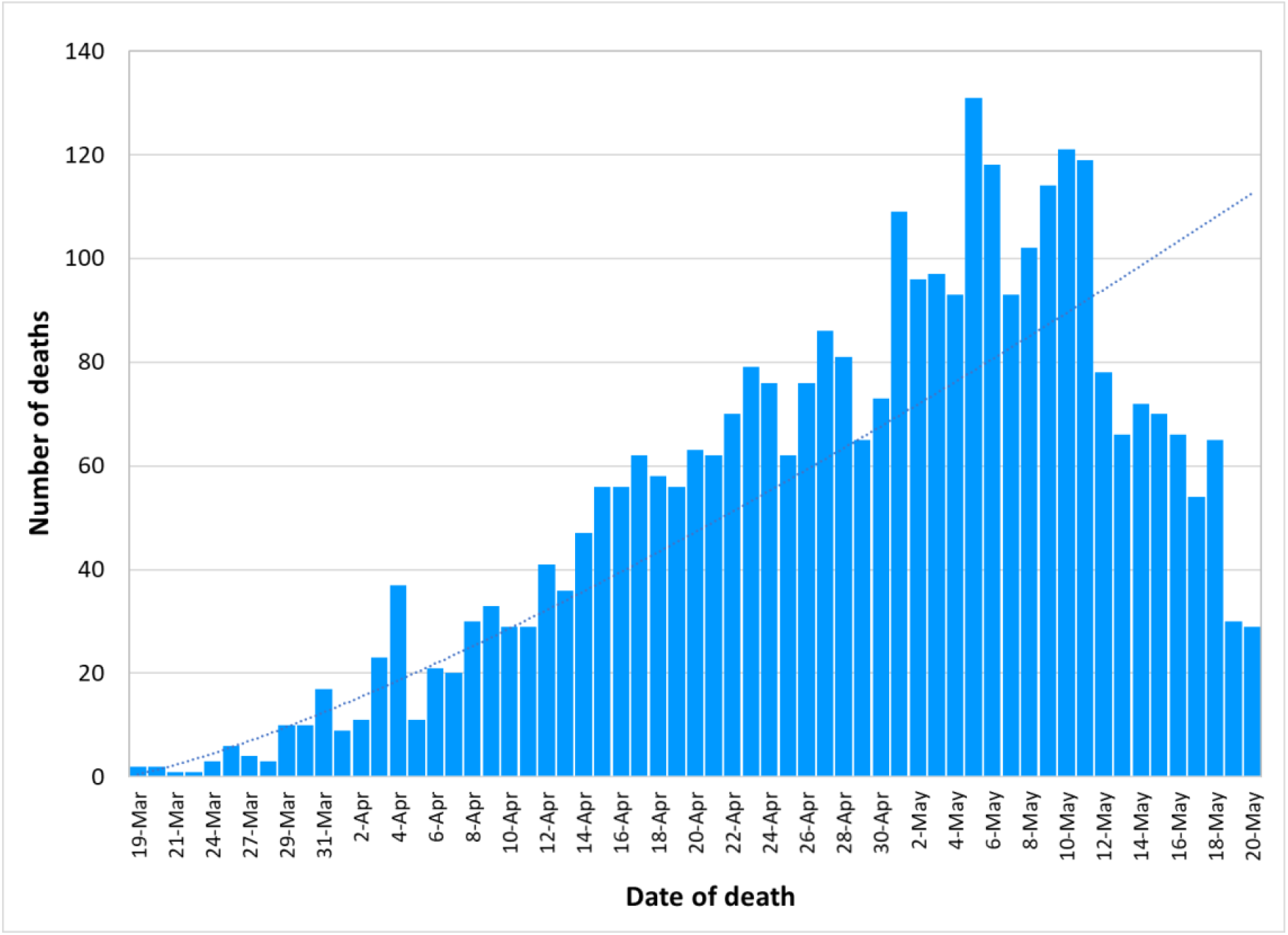
Trend in the number of deaths confirmed by COVID-19 in Peru as of May 22, 2020.

The highest frequency of confirmed deaths from COVID-19 corresponded to males (71.8%), adults over 60 years of age or older (68.3%), and people from the department of Lima (41.8%), observing that 21.8% had some comorbidity. In 81.3% of cases, the death occurred in a public hospital; while, 16.0% died at home, shelter, penitentiary institution, public road, or in transit to a hospital. This is shown in table 1.

**TABLE 1:** Epidemiological characteristics of the deaths confirmed by COVID-19 in Peru.

| CHARACTERISTIC | FREQUENCY | % |
| --- | --- | --- |
| Life stage |  |  |
| Child and adolescent | 19 | 0.6 |
| Young adult | 27 | 0.8 |
| Adult | 982 | 30.3 |
| Elderly | 2216 | 68.3 |
| Gender |  |  |
| Male | 2330 | 71.8 |
| Female | 914 | 28.2 |
| Place of residence |  |  |
| Lima | 1356 | 41.8 |
| Rest of the coast | 1224 | 37.7 |
| Andean region | 259 | 8.0 |
| Amazon | 405 | 12.5 |
| Place of occurrence of the death |  |  |
| Public hospital | 2604 | 81.3 |
| Private clinic | 74 | 2.3 |
| Home, accommodation, shelter, public road, transit to the hospital | 456 | 16.0 |
| Penitentiary institution | 14 | 0.4 |
| Diagnosis of comorbidities |  |  |
| Yes | 1034 | 31.1 |
| No | 2210 | 68.1 |
| Diagnosis with laboratory tests |  |  |
| RT-PCR | 1084 | 33.3 |
| Rapid test IgM detection / IgG | 1857 | 57.1 |
| Both tests | 314 | 9.6 |

## Analysis of pre-hospital and hospital times

### Pre-hospital time

This time averaged was 7.13 ± 4.14 days, observing cases that were hospitalized on the first day of the onset of symptoms up to cases that did so on day 21. The Kruskal Wallis test showed that there was a statistical difference in the pre-hospital time according to the age group of so that adults (Dunn-Bonferroni test; p = 0.001) and elders (Dunn-Bonferroni test; p = 0.002) had a significantly longer time to arrive at the hospital from the onset of symptoms. Likewise, women arrived at the hospital in a significantly shorter time than men (U Mann Withney test; p = 0.037). Other variables such as the region of origin and the health provider institution did not significantly affect pre-hospital time (Table 2).

**TABLE 2:** Comparison of means of pre-hospital and hospital time (Days) with epidemiological characteristics of deaths from COVID-19 in Peru.

| Characteristic | Pre-hospital time (days) |  | Hospital time (days) |  |
| --- | --- | --- | --- | --- |
| | Mean ± standard deviation (median) | p-value | Mean ± standard deviation (median) | p-value |
| Age Group |  |  |  |  |
| Child and adolescent $\Psi$ | 2.90 ± 1.73 (3.00) | <0.001 | 8.36 ± 9.43 (5.00) | <0.001 |
| Young | 6.53 ± 6.72 (5.00) | | 4.58 ± 3.73 (3.00) | |
| Adult | 7.29 ± 3.87 * (7.00) | | 7.62 ± 6.42 (6.00) | |
| Elderly | 7.18 ± 4.29 * (7.00) | | 6.34 ± 5.22μ (5.00) | |
| Gender |  |  |  |  |
| Male $\Psi$ | 7.30 ± 4.17 (7.00) | 0.037 | 6.96 ± 5.67 (5.00) | <0.001 |
| Female | 6.91 ± 4.19 (6.00) | | 6.12 ± 5.59 (4.00) | |
| Region of origin |  |  |  |  |
| Lima $\Psi$ | 7.23 ± 4.38 (7.00) | 0.203 | 7.77 ± 6.26 (6.00) | <0.001 |
| Rest of the coast | 7.27 ± 4.30 (7.00) | | 5.59 ± 4.89 * (4.00) | |
| Andean region | 6.47 ± 3.33 (6.00) | | 6.41 ± 5.08 * (5.00) | |
| Amazon | 7.42 ± 3.73 (7.00) | | 6.52 ± 4.94 (5.00) | |
| Health Care Provider |  |  |  |  |
| Public hospital of the MINSA $\Psi$ | 7.00 ± 4.11 (6.00) | 0.344 | 6.20 ± 5.30 (5.00) | <0.001 |
| EsSalud public hospital | 7.32 ± 4.22 (7.00) | | 6.62 ± 5.51 (5.00) | |
| Hospital of PNP/FF.AA | 6.76 ± 3.94 (7.00) | | 8.25 ± 6.18 * (6.00) | |
| Private clinics | 7.27 ± 4.58 (7.00) | | 9.70 ± 8.00 * $\phi$ (8.00) | |
| Comorbidities |  |  |  |  |
| Yes | 7.06 ± 4.21 (7.00) | 0.247 | 6.77 ± 5.83 (5.00) | 0.797 |
| NoΨ | 7.27 ± 4.16 (7.00) | | 6.71 ± 5.57 (5.00) | |

$\Psi$ Reference category, MINSA: Ministry of Health, EsSalud: Insurance Social of Peru
\* Statistically significant difference compared to the reference category
$\mu$ Statistically significant difference with the adult group
$\phi$ Significant difference with EsSalud

### Hospitalized time

The time averaged was 6.54 ± 5.51 days; thus, some people died on the first day of admission to the hospital up to those who died 38 days after admission after a prolonged stay that included admission to an intensive care unit and mechanical ventilation. The Kruskal Wallis test showed that there was a statistically significant difference in hospital time according to age group, with significantly shorter times in elders compared to adults (Dunn-Bonferroni test; p <0.001), which indicates complications or they died very quickly once they entered the hospital. A similar situation was observed in women that had complications and died in less time than men (Table 2).

There was a statistically significant difference in hospital time according to the region of origin (p <0.001), observing in the post hoc analysis a significantly shorter hospital time in the other departments of the coast (Dunn-Bonferroni test; p <0.001) and the Andean region (Test Dunn-Bonferroni; p = 0.014) compared to Lima (Table 2).

In the case of health care institutions, it is observed that the cases treated in private clinics (Dunn-Bonferroni test; p = 0.001) and hospitals of the National Police and Armed Forces (PNP / FF: AA) patients survived longer time than the people who were treated at MINSA hospitals (Dunn-Bonferroni test; p = 0.032). Hospital time was longer in EsSalud hospitals; however, this difference was not significant (Table 2).

**FIGURE 2:**
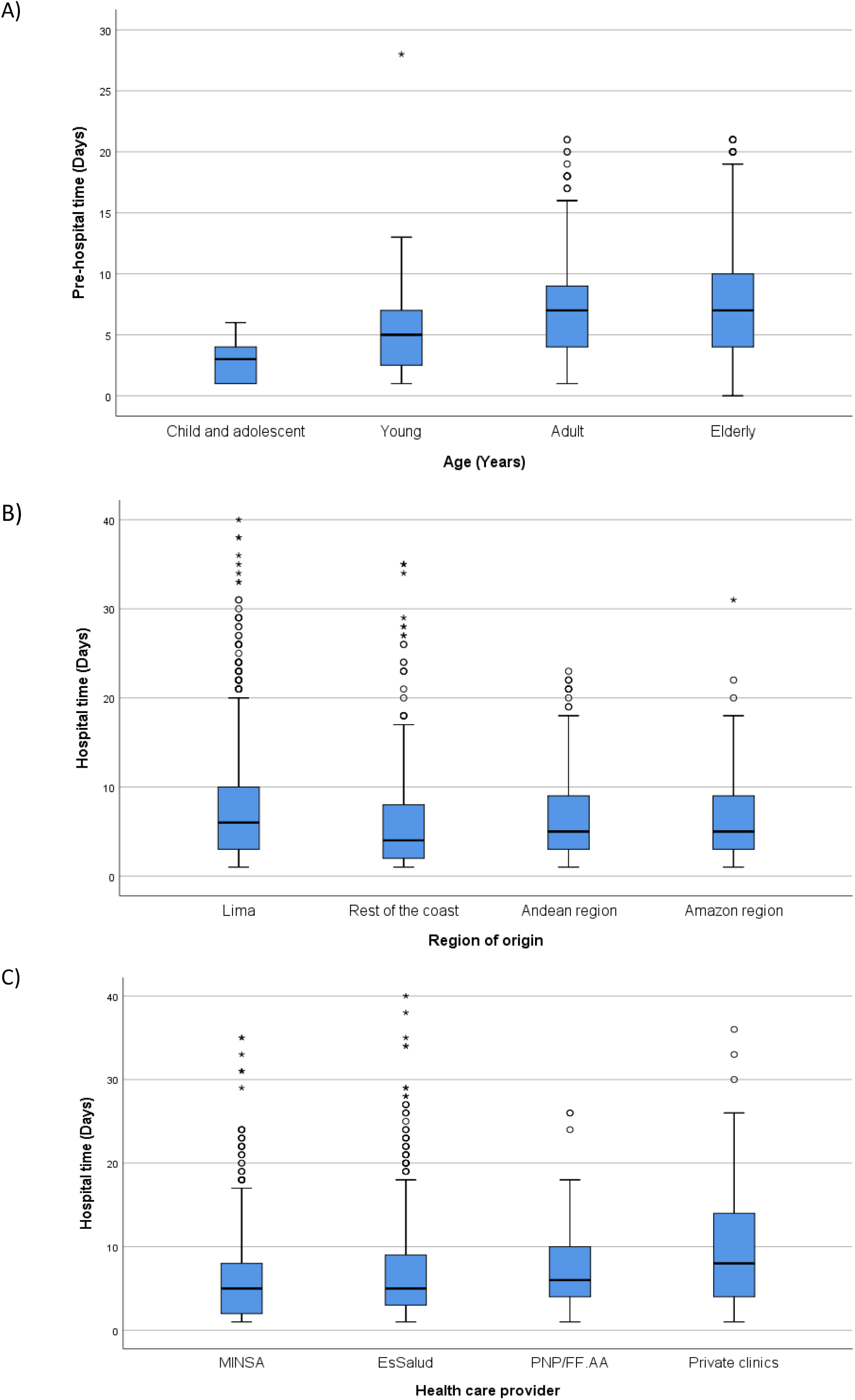
A) Comparison of the pre-hospital time of deaths from COVID-19 according to age group. B) Comparison of hospitalized time (days) of cases of COVID-19 death by region of residence and by health care provider (C).

## DISCUSSION

This research shows that, almost three months after the first confirmed case of COVID-19 in Peru, deaths from COVID-19 were more frequent in males, elders, residents of the department of Lima, and the rest of the Peruvian coast. A third of the deceased had comorbidities; likewise, a considerable fraction died at home, shelter, penitentiary institution, public road or in transit to a hospital. Pre-hospital time is affected by age group and gender; while, hospital time is also influenced by the region of origin and the health provider.

Deaths from COVID-19 were more frequent in elders^17^ which is similar to the study reported by Kang^18^ who finds an exponential increase in the death rate from COVID-19 as age increases, regardless of the geographical region. The greater vulnerability in elders could also be explained by changes that occur in the immune system due to aging and that affect innate and adaptive immunity (Immunosenescence)^17-19^. Studies conducted in China show that the age-related pattern of death from COVID-19 differs from other respiratory viruses, where the severity pattern is often described as a U-shaped curve, with morbidity and mortality concentrated in groups of extreme age (young children and the elderly) unlike COVID-19 which affects adults and is concentrated in elders.^18,20^

In the present series of cases, it is observed that seven out of every ten deaths were men, which could be explained by the greater severity of the disease in them^21,22^. On the other hand, it was observed that a third of deaths from COVID-19 were associated with comorbidity, which could be explained by the greater severity and speed of the clinical picture observed in people with comorbidities that frequently lead to death.^23,24^

Regarding the place of occurrence, 16% of the deaths during the study period were out-of-hospital. Casualty occurred at the patients’ home, accommodation or nursing home, shelter, penitentiary institution, on the public road or in transit to a hospital. The proportion of out-of-hospital deaths is considerably higher than that observed on March 31, 2020, in which 0.2% of deaths confirmed by COVID-19 were out-of-hospital^25^ and on April 30, 2020, where the proportion of out-of-hospital deaths accounted for 8.2 % of total confirmed deaths^26^. This could be related to the collapse of health services, observing that the number of hospital beds, ICU beds, medical specialists, and health professionals available was largely insufficient^27,28^. The high proportion of out-of-hospital deaths that occurred in Peru, as well as in other Latin American countries, reflects the great impact of COVID-19 on health systems, as well as the inequity in the population’s access to health services, particularly in countries with fragmented health systems like Peru.^29-31^

Other aspects that would explain the high proportion of home fatalities were self-medication and insufficient oxygen supply. At first, a significant fraction of the population preferred to self-medicate with hydroxychloroquine, azithromycin, ivermectin, and even chlorine dioxide instead of going to a hospital and causing death due to the rapid evolution of the disease. Later, the supply of oxygen proved to be largely insufficient in public health facilities in the interior of the country as they did not have oxygen plants, which led people to decide to manage their family members at home, acquiring oxygen from private providers combined with the exaggerated increase in the cost of this service, so many families could not afford it. It should be noted that a study carried out in a hospital in Lima^32^ found that oxygen saturation less than 85% was the main predictor of death in people with COVID-19 pneumonia.

Pre-hospital care through rapid response teams, clinical follow-up teams, or medical care in first-level care facilities is essential for the identification, isolation, and treatment of cases, as well as for the early referral of cases to hospitals in case of pneumonia, rapid evolution of the clinical picture or presence of complications^33^. Our research shows that in the deaths studied by COVID-19 there was a significant delay in going to a hospital in adults and elders when this time is compared with that presented by children and adolescents who were taken or referred to a hospital in a very short time. A similar situation occurred with men compared to women who sought care in less time. Contrary to what might be thought, people with a diagnosis of comorbidities did not go to health services more quickly compared to those who did not present comorbidities, which could be related to an inadequate perception of low risk of serious disease similar to the study by Chan reported in China^34^.

On the other hand, hospital time was shorter in the departments of the coast and the Andean region, which indicates that the disease evolved very rapidly and cases died more quickly than in the department of Lima, which has also been documented in countries such as Ecuador, which experienced greater difficulties in cities located on the coast such as Guayaquil^35,36^. This could be due to the delay in the arrival of the cases to health services, as well as the lower resolution capacity of the hospitals in these departments compared to Lima.

Private clinics and hospitals of the PNP / FF.AA showed longer survival time of the cases, in contrast to the establishments of the Ministry of Health and Social Security, which faced high demand for cases that far exceeded the supply provided. Countries like China that were more efficient in improving their response capacity in the short term (Increase in the number of hospitals, the number of hospital beds, beds in intensive care units, and specialized health personnel) presented better results in the management of hospitalized cases. with the consequent reduction in the number of deaths.^37^

Within the limitations, it must be considered that our study was carried out using secondary sources, so it is possible that there are quality problems and underreporting to some degree; however, the fact of having considered several sources of information, as well as the verification and investigation of each death, would partially offset these limitations. Studies carried out in Italy^8^ show that at the peak of the epidemic, out-of-hospital deaths escaped more frequently from the official COVID19 registries, particularly deaths at home or those that occurred in nursing homes.

Another limitation is that the molecular RT-PCR test is not considered a laboratory test in all cases, but rather a significant fraction of cases were diagnosed using rapid tests. Nevertheless, it should be considered that the vast majority of them presented a clinical picture compatible with coronavirus pneumonia and a significant fraction was hospitalized, for this reason, making it difficult for deaths to respond to another etiology.

In conclusion, deaths from COVID-19 occur mainly in male, elder, residents of Lima, and other coastal departments, with considerable deaths at home, in shelters, on public roads, penitentiary institutions, or in transit to a hospital. Pre-hospital time is affected by age group and gender; while, hospital time is also influenced by the region of origin and the health care provider.

It is important to consider these findings to design and disseminate information aimed at the population as well as to improve the response of health services to guarantee timely care to people affected by COVID-19 that due to the severity of their clinical picture requires hospital care, not only in Peru but also in other countries with a similar epidemiological panorama.

## Data Availability

The databases consulted and the sources are freely accessible and available.

https://www.dge.gob.pe/portal/docs/tools/coronavirus/coronavirus300420.pdf

https://www.dge.gob.pe/portal/docs/tools/coronavirus/coronavirus200520.pdf.

https://www.dge.gob.pe/portal/docs/tools/coronavirus/coronavirus310320.pdf

https://www.dge.gob.pe/portalnuevo/covid-19/covid-cajas/situacion-del-covid-19-en-el-peru/

